# Polygenic risk scores as a marker for epilepsy risk across lifetime and after unspecified seizure events

**DOI:** 10.1101/2023.11.27.23297542

**Authors:** Henrike O. Heyne, Fanny-Dhelia Pajuste, Julian Wanner, Jennifer I. Daniel Onwuchekwa, Reedik Mägi, Aarno Palotie, FinnGen, Estonian Biobank research team, Reetta Kälviainen, Mark J. Daly

## Abstract

A diagnosis of epilepsy has significant consequences for an individual but is often challenging in clinical practice. Novel biomarkers are thus greatly needed. Here, we investigated how common genetic factors (epilepsy polygenic risk scores, [PRSs]) influence epilepsy risk in detailed longitudinal electronic health records (EHRs) of > 360k Finns spanning up to 50 years of individuals’ lifetimes. Individuals with a high genetic generalized epilepsy PRS (PRS_GGE_) in FinnGen had an increased risk for genetic generalized epilepsy (GGE) (hazard ratio [HR] 1.55 per PRS_GGE_ standard deviation [SD]) across their lifetime and after unspecified seizure events. Effect sizes of epilepsy PRSs were comparable to effect sizes in clinically curated data supporting our EHR-derived epilepsy diagnoses. Within 10 years after an unspecified seizure, the GGE rate was 37% when PRS_GGE_ > 2 SD compared to 5.6% when PRS_GGE_ < –2 SD. The effect of PRS_GGE_ was even larger on GGE subtypes of idiopathic generalized epilepsy (IGE) (HR 2.1 per SD PRS_GGE_). We further report significantly larger effects of PRS_GGE_ on epilepsy in females and in younger age groups. Analogously, we found significant but more modest focal epilepsy PRS burden associated with non-acquired focal epilepsy (NAFE). We found PRS_GGE_ specifically associated with GGE in comparison with >2000 independent diseases while PRS_NAFE_ was also associated with other diseases than NAFE such as back pain. Here, we show that epilepsy specific PRSs have good discriminative ability after a first seizure event i.e. in circumstances where the prior probability of epilepsy is high outlining a potential to serve as biomarkers for an epilepsy diagnosis.

## Introduction

Epilepsy is a serious neurological disorder characterized by unprovoked seizures, which affects up to 1% of individuals worldwide (WHO, 2019), with children and the elderly being particularly affected. Although epilepsy can be caused by acquired conditions such as stroke, tumor or head injury, most cases (ca. 70–80%) are due to genetic influences ^1^, including rare and common genetic variants. Diagnosing epilepsy is often challenging ^2–4^ and multiple individuals are initially misdiagnosed ^5^. An epilepsy diagnosis is potentially lifesaving with a 3× elevated mortality risk in epilepsy (WHO, 2019). Epilepsy–related deaths can be prevented by antiseizure medication (ASM) which however often have adverse effects ^6^. Thus, correct epilepsy diagnosis is crucial, but the most widely-used diagnostic tool in epilepsy, the electroencephalogram (EEG), has quite variable sensitivity and specificity in different clinical settings ranging about 17%-58% and 70–98%, respectively ^7,8^ and moderate inter-rater agreement ^9^, illustrating a need for additional biomarkers ^3^. Due to the great importance and challenge specific ‘first seizure clinics’ are solely dedicated to investigating an epilepsy diagnosis after a newly onset seizure ^2^. Until 2014, epilepsy was defined as having two unprovoked seizures >24 h apart by the International League Against Epilepsy ^10^. This definition was then extended to the following additional scenarios of having 2) a diagnosis of an epilepsy syndrome or 3) one unprovoked seizure and a probability of further seizures with a recurrence risk of at least 60% over the next 10 years ^10^.

The common epilepsies can be broadly categorized into genetic generalized (GGE) and non-acquired focal epilepsies (NAFE), where the latter originate from a particular brain area ^10^. First-degree relatives of patients with GGE had an 8.3-fold increased risk of developing GGE while first-degree relatives of patients with NAFE had a 2.5-fold increased risk to develop NAFE, compared to the general population, respectively ^11^. In agreement, the SNP-heritability (i.e., the variance of GGE attributed to common genetic variants) is approximately 30-40% ^12,13^ which is relatively high compared to other common diseases ^14^. The same measure is more moderate for NAFE with SNP-heritability of about 9-16% ^12,13^. Previous genome-wide association studies have shown that common variants contribute more substantially to the more common forms of epilepsy ^12^. There is only a modest burden of ultra-rare genetic variants in GGE and NAFE; rare variants likely contribute only a small fraction towards their heritability ^15^ and there are few Mendelian disease genes exclusively associated with them ^16^.

Recent research has shown that common genetic variants with small effects on specific diseases can be combined into “polygenic” risk scores (PRSs) with high disease-specific PRSs conferring comparable risk as rare monogenic variants ^17^. Thus, interest in PRSs is growing as a potential clinically important diagnostic tool ^18–22^. It was recently shown that individuals with epilepsy had a significantly higher epilepsy PRS compared to unaffected controls ^23,24^. However, investigation how epilepsy PRSs may predict epilepsy risk in specific clinical scenarios has so far been lacking. Here, we thus investigate how epilepsy PRSs can stratify epilepsy risk across lifetime and after unspecified seizure events.

## Results

### Electronic health records accurately represent epilepsy diagnoses

We investigated epilepsy PRSs in detailed longitudinal electronic health records (EHR) from the FinnGen project ^25,26^ using FinnGen data freeze R8 (n=342,378, 190,837 females) and the Estonian biobank ^27^ as a validation cohort (for further study sample characteristics see Table 1). We further explored the Bio*Me* cohort ^28^ with regards to Non-European ancestries. Our phenotype data was derived from ICD codes and ASM purchases and reimbursements of official state registries spanning up to 50 years. We defined non-acquired focal epilepsy (=NAFE) by having ≥2 NAFE specific ICD codes and genetic generalized epilepsy (=GGE) by having ≥2 GGE specific ICD codes, respectively. Additionally, we require ≥2 ASM purchases for a NAFE or GGE category (Supplementary Figure 1). Of 606 individuals with diagnosis codes of both NAFE and GGE we could categorize 518 as either GGE or NAFE also using reimbursement data. Further details on epilepsy case definitions can be found in Supplementary Tables 1 and 2 and Methods. Sample numbers are given in Table 1. We also investigated 131 individuals with ≥2 traditional idiopathic generalized epilepsy diagnoses (=IGE) ^29^ i.e. Childhood/Juvenile Absence Epilepsy (ICD 40.33/35), Juvenile Myoclonic Epilepsy (ICD 40.36) and Generalized Tonic–Clonic Seizures Alone (ICD 40.35). Individuals’ age at first epilepsy diagnosis was in line with known age of onset of respective IGE syndromes supporting our EHR-derived diagnoses (Supplementary Figure 2.)

**Table 1.** Descriptive Statistics of main study and replication cohort. FinnGen; FinnGen study (Finland), EstBB; Estonian biobank (Estonia), repl; replication cohort. All ages and times are given in years.

| **Study Sample Characteristics** | **FinnGen** | **EstBB (repl.)** |
| --- | --- | --- |
| Total participants | 342,378 | 210,382 |
| Female (%) | 53.6 | 65.4 |
| Control (n) | 273,974 | 164,346 |
| GGE (n) | 622 | 275 |
| NAFE (n) | 3,127 | 1,233 |
| one unspecified seizure (n) | 1,806 | NA |
| Age Control (mean±SD) | 57.9 ± 18.6 | 49.1 ± 16.3 |
| Age GGE (mean±SD) | 43.4 ± 19.9 | 42.3 ± 16.0 |
| Age NAFE (mean±SD) | 57.1 ± 19.7 | 53.2 ± 17.4 |
| Age at first GGE diagnosis (mean±SD) | 25.3 ± 18.2 | 29.5 ± 16.3 |
| Age at first NAFE diagnosis (mean±SD) | 43.6 ± 22.5 | 41.7 ± 19.6 |
| Age at unspecified seizure event (mean±SD) | 36.6 ± 28.5 | NA |
| Follow-up time after unsp. seizure (mean±SD) | 14.8 ± 10.7 | NA |
| Ancestry (continental) | European | European |

### Epilepsy PRS most elevated in GGE, specifically IGE, with same effect sizes as in clinically curated epilepsy cohorts

We then calculated epilepsy PRSs to determine individuals’ genetic burden for epilepsy. Here, we used the International League Against Epilepsy (ILAE) GWAS 2018 summary statistics ^12^ as discovery data, i.e. to determine which genetic variants increase or decrease epilepsy risk. We then summed 1000s of genetic risk/protective variants for epilepsy with individually small effects into a single epilepsy PRS per individual. Here, we constructed separate focal epilepsy PRS (PRS_NAFE_) and generalized epilepsy PRS (PRS_GGE_). We found a significant elevation of PRS_GGE_ in individuals with GGE (Figure 1A) which was particularly pronounced in IGE (see also next paragraph and Figure 1B). We also found a significant elevation of PRS_NAFE_ in 2,310 individuals with NAFE (Figure 1C), but no significant elevation of PRS_GGE_ in individuals with unspecified seizures (Figure 1D). Similarly, we found a high correlation (Pearson’s correlation coefficient =0.9, p-value = 1×10^−7^) between PRS_GGE_ decile and GGE prevalence in our data (Figure 2). Overall, we can thus confirm previous studies that used PRS as a marker for genetic liability of common epilepsy types. Importantly, we find very similar respective effect sizes of PRS_GGE_ and PRS_NAFE_ on GGE and NAFE (see Supplementary Table 3) as reported in previous cohorts from Epi25 or the Cleveland Clinic ^23^. We thus consider it very likely that the epilepsy phenotypes in our biobank data are comparable to the phenotypes curated according to clinical criteria in these cohorts.

**Figure 1.**
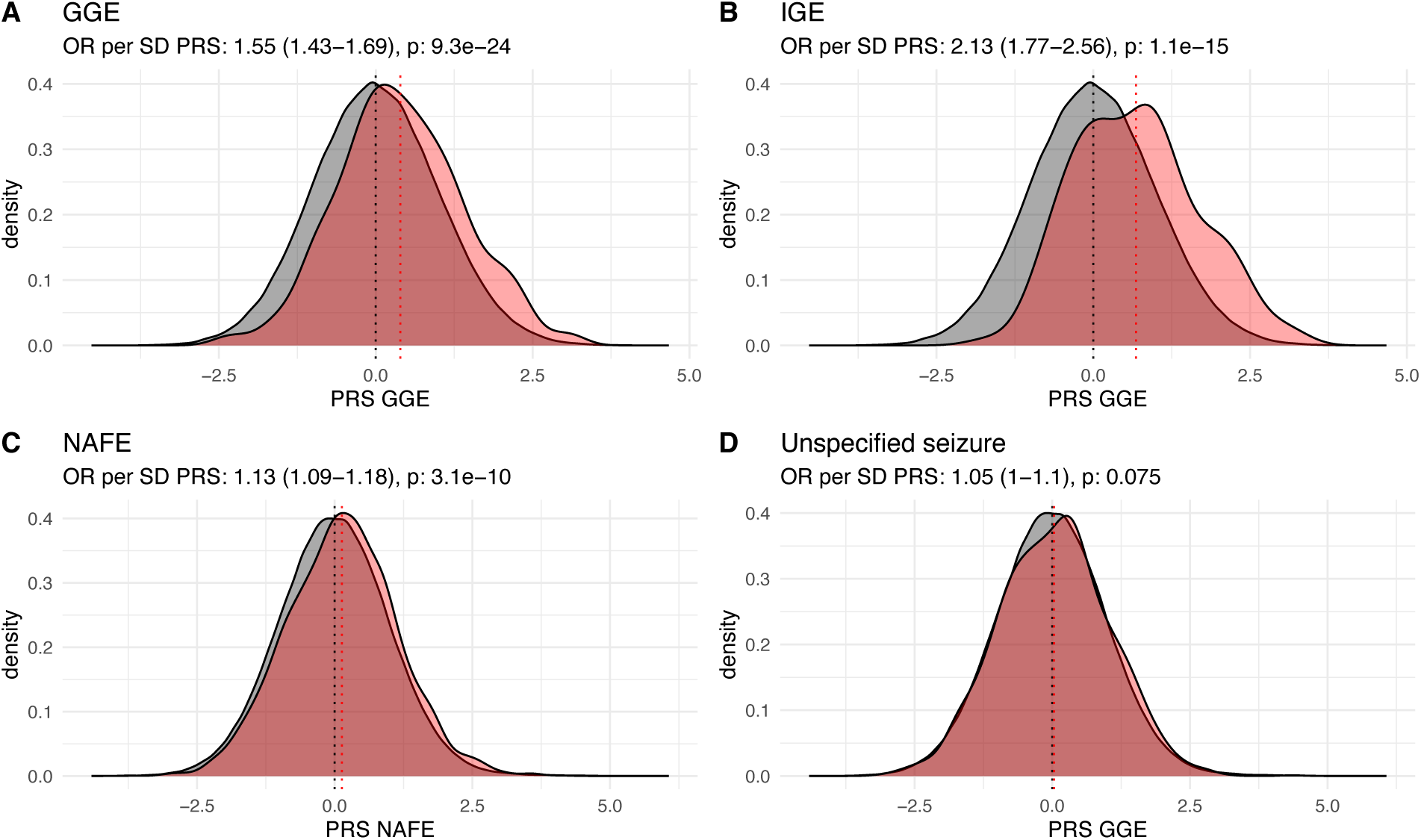
Epilepsy PRS of epilepsy cases (red) compared to population controls (grey) (n=273,974) (density curves). A) PRS_GGE_ of GGE (n=622) compared to controls B) PRS_GGE_ of IGE (n=131) compared to controls C) PRS_NAFE_ of NAFE (n=3,127) compared to controls D) PRS_GGE_ of unspecified seizure without epilepsy (n=1,806) compared to controls. On top of each panel: odds ratios (95%-confidence intervals in brackets) and p-values of standard deviation increase of epilepsy PRS on case versus control status. Mean of the PRS distributions are shown as vertical dotted lines. Method: logistic regression. OR; odds ratio, p; p-value.

**Figure 2.**
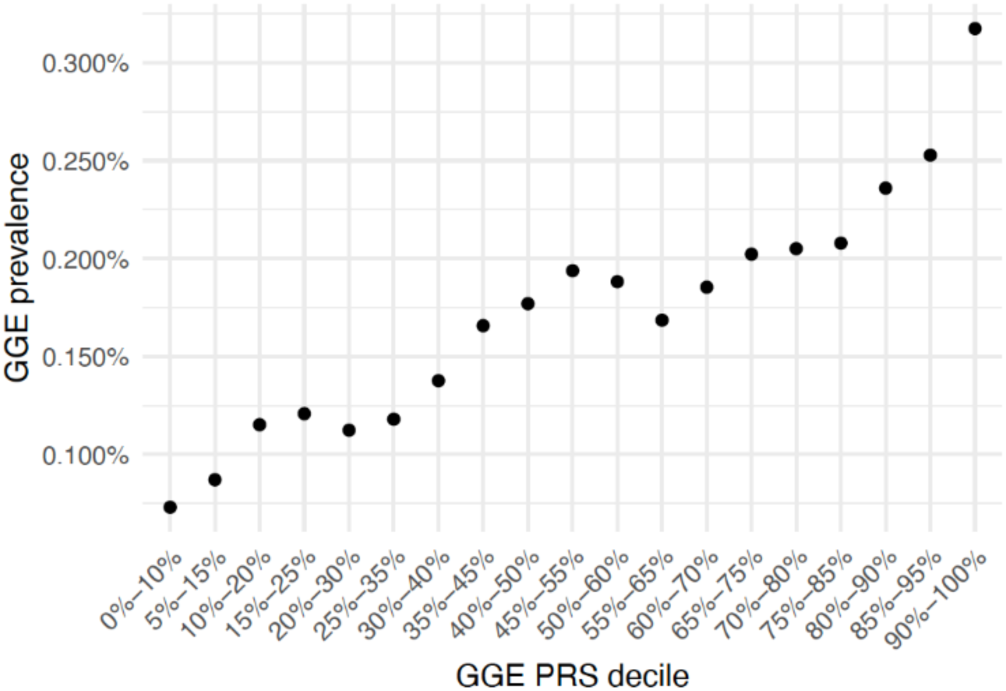
Correlation of PRS_GGE_ decile and GGE prevalence. The bins of adjacent PRS deciles are overlapping, as labelled.

### High PRS is associated with epilepsy across lifetime and after unspecified seizure events

We next investigated the effect of epilepsy PRS on epilepsy rates across lifetime, separately for PRS_GGE_ and PRS_NAFE_. We stratified our cohort into bins of epilepsy PRS standard deviations (SD) (Figure 3) and compared the cumulative epilepsy incidence in each SD bin to the rest of the cohort (for increased power). Individuals with a PRS_GGE_ > 2 SD (ca. 2% of the cohort) had a more than tripled lifetime risk to develop GGE than the rest of the cohort (Hazard ratio [HR]: 3.2, Confidence Interval [CI]: 2.1-4.5, p-value: 2×10^−9^, method: cox proportional hazard model ^30^ [coxph], Figure 3, panel A). The epilepsy risk decreased proportionally with the decreasing PRS_GGE_ SD bin. Overall, the HR increased by 1.6 per increased SD of PRS_GGE_ (95%-CI 1.4-1.7, p-value=7×10^−24^). When restricting to IGE the HR per PRS_GGE_ SD was 2.1 (95%-CI 1.8-2.6, p-value 1×10^−15^). Individuals with a PRS_GGE_ > 1 SD had a HR of 29.4 for IGE compared to those with PRS_GGE_ < –1 SD (95%-CI 7-124, p-value 4×10^−6^, IGE rate in PRS_GGE_ < –1 SD: 2/43,977, IGE rate in PRS_GGE_ > 1 SD: 45/43,827). PRS discriminated GGE cases versus controls with a concordance index (C-index) of 0.61 (95%-CI 0.56-0.65) adjusting for the same covariates (birth year, sex, batch and PCs). Overall, we thus showed PRS_GGE_ as a significant biomarker for lifetime epilepsy risk.

**Figure 3.**
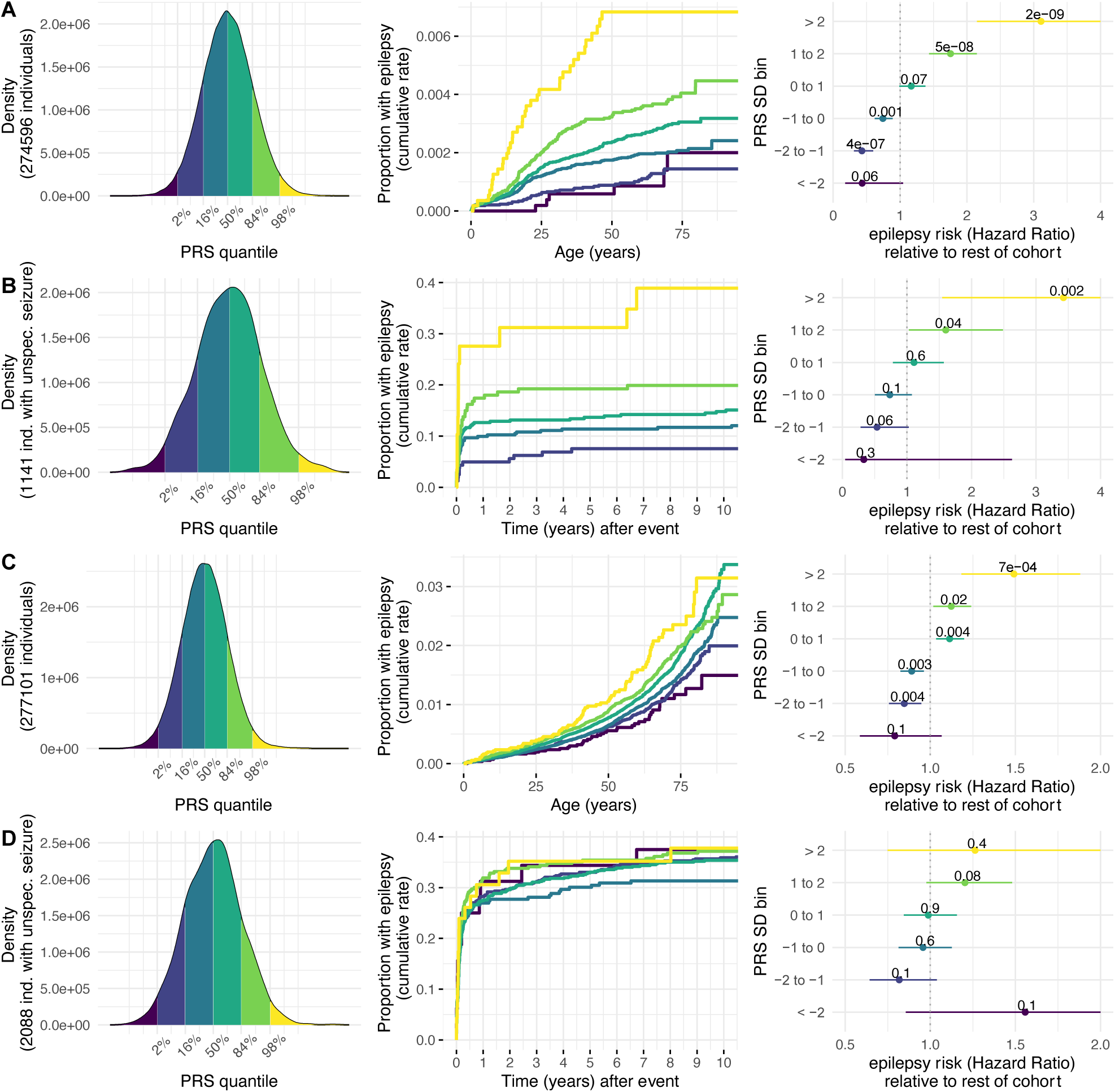
Epilepsy PRS as a marker for epilepsy risk across lifetime and after unspecified seizure events. Panel A) GGE across lifetime, B) GGE after an unspecified seizure, C) NAFE across lifetime, D) NAFE after an unspecified seizure. In panels A/B, we investigate PRS_GGE_, in panels C/D, PRS_NAFE_. In each panel, on the left are density curves that display how samples are partitioned into six bins of PRS standard deviations. Survival curves in the middle give the cumulative epilepsy incidence (y-axis) across time (x-axis [years]) stratified for epilepsy PRS bins. In panel B, the two lowest PRS bins are fused in the survival curve as epilepsy case counts were too low. The rightmost figures show epilepsy risk of each epilepsy PRS bin compared with the rest of the cohort (forest plots). Here, the point estimates represent hazard ratios, error bars show the 95%-confidence intervals (CIs). For clarity, the CIs are capped at 4 (panels A and B) and 2 (panels C and D).

However, the absolute risk to develop epilepsy is small across lifetime (<1%, see Figure 3A/C), even for individuals with high epilepsy PRS. Lifetime risk prediction is thus less clinically meaningful. When considering the subset of individuals that were diagnosed with an unspecified seizure corresponding to ICD code R56.8/7803A at an age < 40 years their absolute risk for GGE increased compared to baseline (Figure 3B). Within 10 years after the unspecified seizure, the GGE rate reached 37% in > 2 SD PRS_GGE_ compared to 5.6% in < –2 SD PRS_GGE_ (or 23% in > 1 SD PRS_GGE_ versus 7.4% in < –1 SD PRS_GGE_). PRS_GGE_ affected relative epilepsy risk similarly after an unspecified seizure (HR per PRS_GGE_ SD: 1.5, 95%-CI: 1.3-1.8, p-value=1×10^−5^, C-index 0.60, 95%-CI 0.51-0.68) as across lifetime. Similarly, PRS_NAFE_ had a significant but more modest effect on NAFE cumulative lifetime incidence (HR per PRS_NAFE_ SD: 1.13, 95%-CI: 1.09-1.17, p-value=3×10^−10^) and after unspecified seizure (HR per PRS_NAFE_ SD: 1.07, 95%-CI: 1.02-1.2, p-value=0.01), in line with a lower heritability of focal epilepsy (Figure 3C/D).

We replicated analyses of PRS_GGE_ effects across lifetime in the Estonian biobank ^27^ (Estonia, European ancestry, Supplementary Figure 3) obtaining very similar estimates (see Table 2) and thus validating our results. We further explored the effects of PRS_GGE_ in individuals with diverse ancestries in the Bio*Me* biobank (New York City, USA, Supplementary Note, Supplementary Figure 4). While the effect of PRS_GGE_ in Bio*Me* followed similar trends, our analyses were underpowered and thus did not reach significance. Further analyses are those needed to investigate the portability of epilepsy PRS effects to other ancestry groups.

**Table 2.**
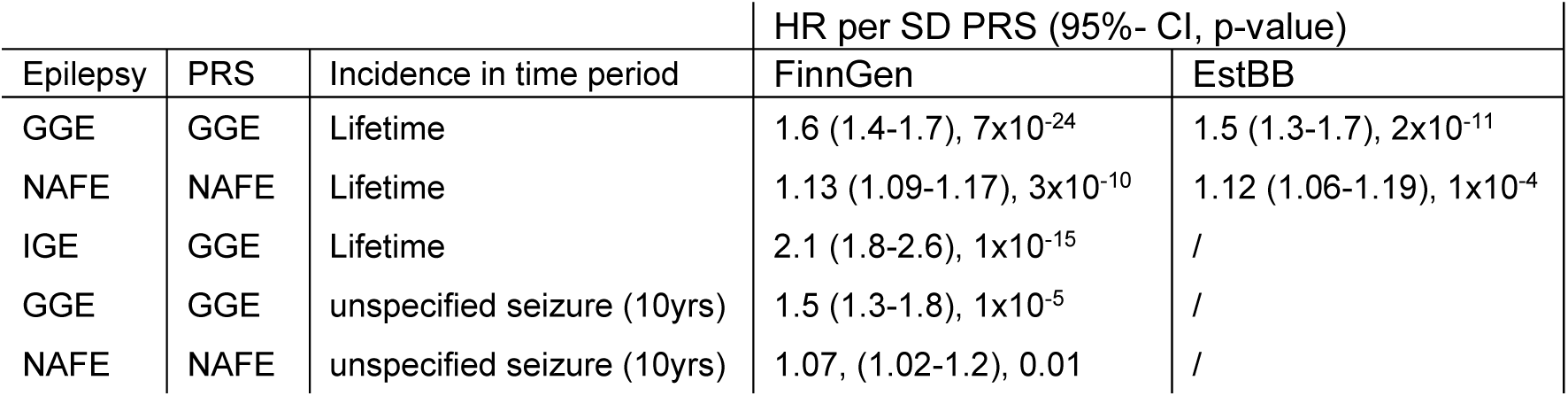
Effects of PRS on epilepsy risk in biobanks FinnGen and Estonian biobank. Epilepsy risks are given as hazard ratios (HR) for epilepsy per SD increase of epilepsy PRS. Method: Cox proportional hazards model. CI; confidence interval, FinnGen; FinnGen study (Finland), EstBB; Estonian biobank (Estonia), unspecified seizure (10yrs); 10 years after an unspecified seizure

| Epilepsy | PRS | Incidence in time period | HR per SD PRS (95%- CI, p-value) |  |
| --- | --- | --- | --- | --- |
|  |  |  | FinnGen | EstBB |
| GGE | GGE | Lifetime | 1.6 (1.4-1.7), $7 \times 10^{-24}$ | 1.5 (1.3-1.7), $2 \times 10^{-11}$ |
| NAFE | NAFE | Lifetime | 1.13 (1.09-1.17), $3 \times 10^{-10}$ | 1.12 (1.06-1.19), $1 \times 10^{-4}$ |
| IGE | GGE | Lifetime | 2.1 (1.8-2.6), $1 \times 10^{-15}$ | / |
| GGE | GGE | unspecified seizure (10yrs) | 1.5 (1.3-1.8), $1 \times 10^{-5}$ | / |
| NAFE | NAFE | unspecified seizure (10yrs) | 1.07, (1.02-1.2), 0.01 | / |

### Effect of age and sex on association of epilepsy PRS with epilepsy

In other diseases than epilepsy, studies previously reported sex-specific PRS effects and larger effects of PRS on disease in earlier age groups ^31^. Thus, we sought to investigate the effect of age at onset and sex on PRS effects on epilepsy. We divided our cohort into quintiles of age at onset and found significant effects of PRS_GGE_ on GGE and of PRS_NAFE_ on NAFE case status in all age bins except GGE onset > 60 years and NAFE onset > 80 years (method logistic regression, see Supplementary Table 4). We found the largest effects of PRS_GGE_ on GGE at onset 0-20 (OR 1.6, 95%-CI 1.4-1.8, p-value 2×10^−13^) and 20-40 years (OR 1.6, 95%-CI 1.4-1.9, p-value 1×10^−9^) and of PRS_NAFE_ on NAFE in the 20-40 (OR 1.15, 95%-CI 1.06-1.24, p-value 7×10^−4^) and 40-60 year (OR 1.13, 95%-CI 1.06-1.22, p-value 4×10^−4^) bins. This is in line with other illnesses ^31^. We next investigated, if the large genetic influences on IGE described in the paragraph above could be explained by a higher proportion of individuals with younger age at epilepsy onset in the IGE group. So within the GGE group, we compared the effect of PRS_GGE_ on IGE versus non-IGE and found a higher effect of PRS_GGE_ on IGE even when accounting for age at first epilepsy diagnosis (OR 1.62, 95%-CI 1.24-2.16, p-value 6×10^−4^).

We then wanted to know whether sex influenced epilepsy PRS effects and found a significant interaction of sex and PRS_GGE_ on GGE (p-value 0.004). So we next investigated the effect of PRS on epilepsy separately for men and women. PRS_GGE_ had an unexpectedly larger influence on lifetime GGE in females (HR per PRS SD: 1.7, 95%-CI 1.5-1.9, p-value=8×10^−23^, n=374) than in males (HR per PRS SD: 1.3, 95%-CI 1.1-1.5, p-value=2×10^−4^, n=218, Supplementary Figure 5). We found no significant interaction of sex and PRS_NAFE_ on NAFE (p-value 0.4) and hence similar effects in males and females on NAFE (HR_male_: 1.1, 95%-CI 1.05-1.16, p-value=2×10^−4^; n=1460, HR_female_: 1.15, 95%-CI 1.09-1.2, p-value=2×10^−7^, n=1574).

### PRS_GGE_ is specifically associated with GGE while PRS_NAFE_ is more heterogeneous

We aimed to investigate the phenotypes associated with a genetic epilepsy liability that are *not* epilepsy, in a hypothesis-free approach to elucidate whether genetic factors influence GGE/NAFE in a disease-specific manner. We thus performed a phenome-wide association study (PheWAS) testing the effect of PRS_GGE_ and PRS_NAFE_ on 2,139 distinct disease phenotypes (method: logistic regression, Figure 4). GGE (labeled as ‘Generalized Epilepsy’) is the only phenotype that is significantly affected by PRS_GGE_ after Bonferroni correction. We thus argue that PRS_GGE_ is very specifically associated with GGE increasing its potential diagnostic utility. While PRS_NAFE_ is expectedly associated with NAFE, multiple other phenotype associations are unexpected. The most significant ones are related to back pain, but also include hypertension, cardiovascular disease and depression medications, with lower significance. We tested the genetic correlation of NAFE and the 19 traits that were significant in our PheWAS (method: LD score regression ^32,33^, Supplementary Figure 6). After multiple testing correction none remained significant. However, phenotypes ‘other anxiety disorders’ (*r*_g_ = 0.54, p-value= 0.02), ‘all anxiety disorders’ (*r*_g_ = 0.44, p-value= 0.02) and ‘depression medications’ (*r*_g_ = 0.33, p-value= 0.04) were genetically correlated with nominal significance.

**Figure 4.**
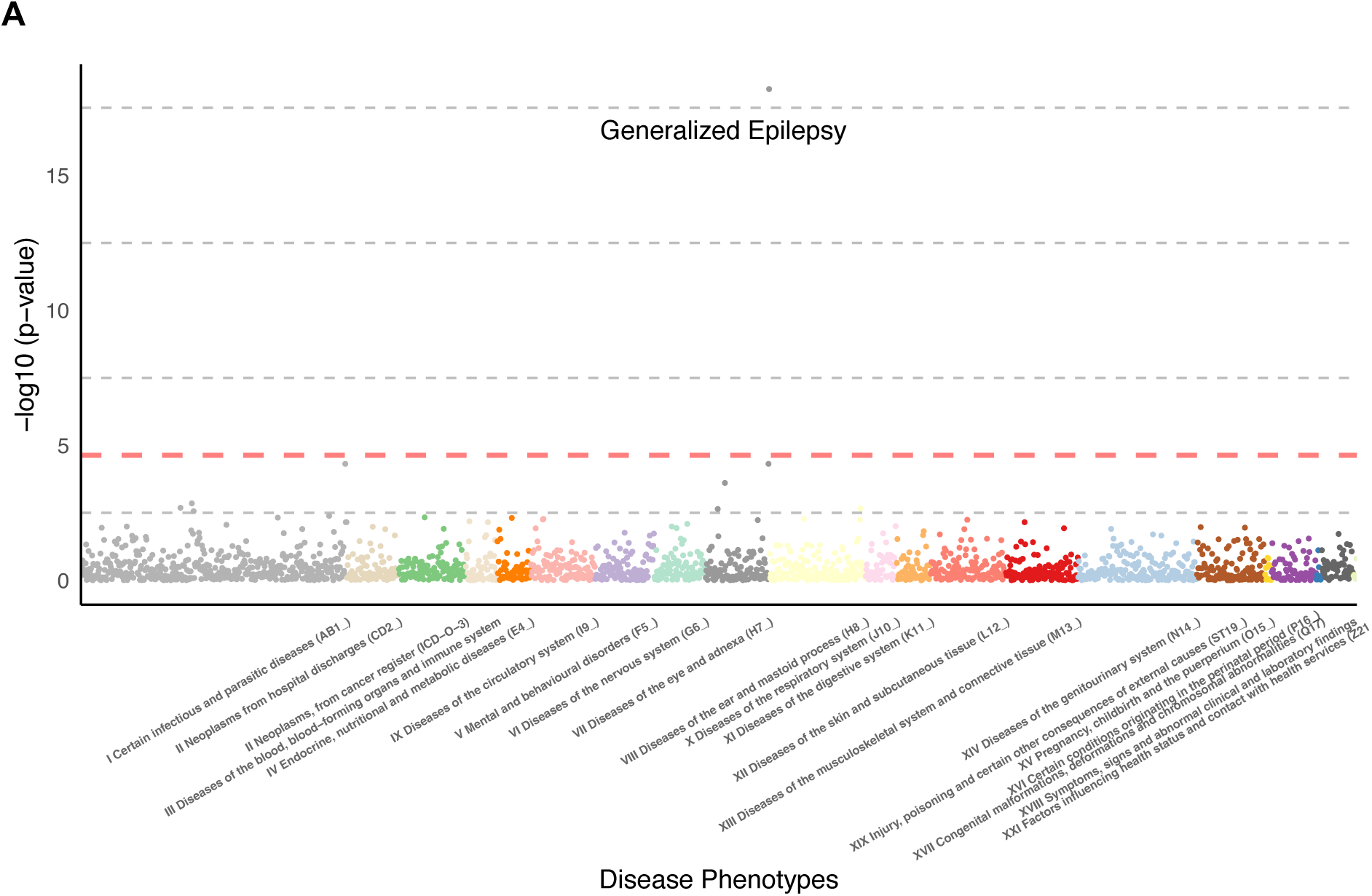
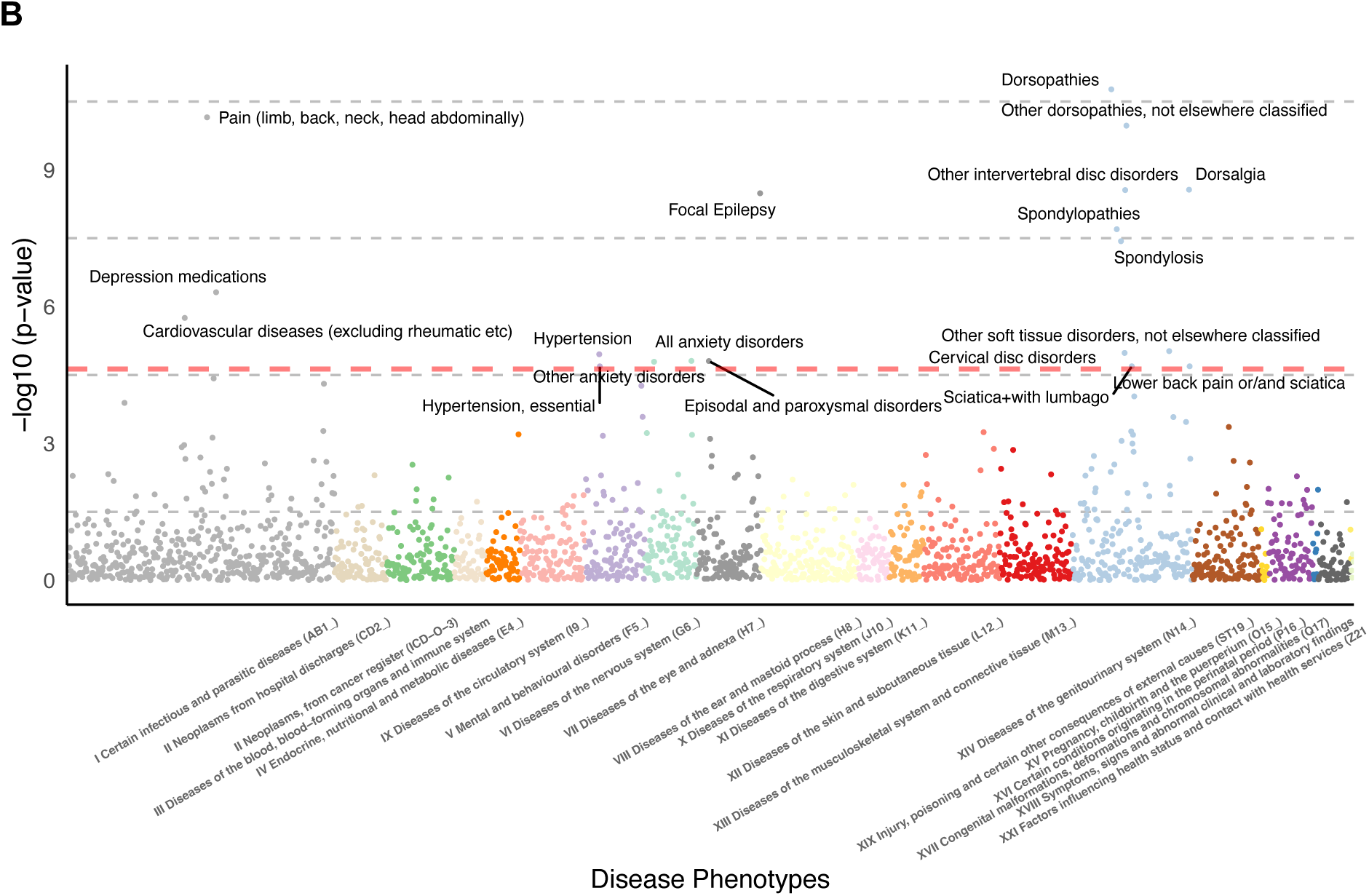
Phenome-wide association study testing the effect of epilepsy PRS on 2139 distinct disease phenotypes in FinnGen. In panel A) we display a PheWAS of PRS_GGE_. GGE, here labeled as ‘Generalized Epilepsy’, is the only phenotype that is significant after Bonferroni correction. In B) we display a PheWAS of PRS_NAFE_. In both panels, the phenotypes are grouped and colored by clinical field (x-axis). The y-axis shows the –log10 p-value of the association (method: logistic regression). The dashed orange horizontal line at p-value 2.3×10^−5^ is the significance threshold after multiple testing correction (Bonferroni).

## Discussion

The diagnosis of epilepsy is an important yet challenging clinical task; thus the need for novel biomarkers remains high. Recent studies demonstrated a genetic burden in the form of an elevated PRS_GGE_ in epilepsy cases versus controls ^23,24^ which we replicate in our data. The effect of PRS_GGE_ has however not been studied outside the case control setting. Here, we investigate the effect of PRS_GGE_ longitudinally; on lifetime epilepsy, on epilepsy after an unspecified seizure event and on 1000s of disease endpoints in other clinical areas.

In this study we could demonstrate that common genetic variants, in the form of PRS_GGE_ have a significant quantitative effect on GGE lifetime cumulative incidence that we could reproduce in several biobanks with hazard ratios of 3-4 for the upper tails of the PRS_GGE_ distribution in line with previous studies ^23^. Prediction is modest, but comparable to performance of models using standard variables such as clinical factors or EEG ^34,35^, so we expect PRSs to have most utility as a supportive but not standalone tool.

We were surprised to find that the effect of PRS_GGE_ on GGE was substantially larger in females than males which was not previously reported. Previous studies reported a higher incidence of GGE in women ^36,37^ which we also observed in our cohort. These could be caused by a different epilepsy susceptibility in males and females mediated by biological or environmental sex-specific factors. This is likely not caused by different pathomechanisms as a recent study found a high correlation of genetic effects on epilepsy in males and females ^13^. Thus, further research is needed to elucidate how genetic factors may differently influence epilepsy between sexes. In our data, the effect of PRS_NAFE_ on NAFE across lifetime was also significant but more modest than for PRS_GGE_ and GGE, in line with other studies.

The effect of PRS_GGE_ on GGE was very specific with no significant effects on other diseases. This should increase its potential utility as a biomarker. Of course, we did not test effects on non-disease phenotypes. Previously, high PRS_GGE_ and high PRS_NAFE_ were both associated with low educational attainment and neuroticism-related personality traits ^38^ which could result from epilepsy or side effects of ASMs or may also be pleiotropic effects. Apart from NAFE, PRS_NAFE_ had effects on other diseases including back pain, which was not previously reported; and anxiety/depression related traits. Here, nominal significant genetic correlations of NAFE with anxiety disorders and depression medications are in line with previous reports in the UK biobank that individuals with high PRS_NAFE_ but without a NAFE diagnosis had more likely experienced anxiety or depression ^38^ pointing to a potential pleiotropic effect.

We see the highest potential clinical utility of epilepsy PRS in patient groups with a high absolute risk to have epilepsy such as after an unspecified seizure event. Current clinical guidelines require at least one unprovoked seizure and at least a 60% chance of a second seizure to diagnose epilepsy ^10^. In a clinical setting, the diagnosis is often not as quantifiable as the definition suggests and heavily dependent on clinical expertise. We find, as an example, that individuals with a PRS_GGE_ >2 SD have a ca. 3× increased risk to be diagnosed with GGE than the rest of the population. This includes the group of individuals with unspecified seizure events who are at elevated risk for a later epilepsy diagnosis. After exclusion of reversible causes for their unspecified seizure, a high PRS_GGE_ could support stratifying groups at risk for a second seizure in conjunction with an EEG while other biomarkers are currently sparse ^3^. Other recent studies suggest that PRS_GGE_ have additional value to the information of family history ^39,40^. Practically, genetic testing is often routinely done in pediatric epilepsy and PRS generation could thus potentially be integrated in an existing workflow. Here, integrating PRS with rare variants could also improve disease prognosis as genetic background has been shown to influence how severely carriers of “monogenic diseases” ^41^ including Dravet syndrome ^42^ are affected. Another advantage is a high cost-effectiveness as PRS can be generated from genotype data that can also be repurposed from other disease areas ^22^.

Our study has several limitations. We have conducted most of our analyses in cohorts with European ancestry. As has been previously described for other diseases, the predictive ability of polygenic risk scores is heavily dependent on genetic ancestry ^43^. While we could replicate our findings in the diverse primarily non-European Bio*Me* cohort across ancestries, sample sizes of individual ancestry groups remained prohibitive. Further studies in diverse populations are thus needed. Another limitation is that our phenotype data is derived from EHRs. We can thus not verify how many epilepsy cases have been confirmed by epileptologists. However, we obtain almost identical PRS effect sizes as in clinical cohorts ^23^, which thus validates our case definitions combining EHR diagnoses with ASM purchase and reimbursement data. The central registry of Finnish EHR data have the unique advantage that reimbursements for ASMs are always based by certificate made by a neurologist.

Our data thus proposes an interesting potential for epilepsy PRS, specifically for PRS_GGE_, as a biomarker for epilepsy risk where it could – combined with clinical markers such as the EEG – improve epilepsy risk prediction. Our data outlines how this could be specifically useful in situations of elevated epilepsy risk such as an unspecified seizure event. Ultimately, this needs to be investigated in a clinical setting.

## Supporting information

Estonian biobank banner

FinnGen banner

## FinnGen

We want to acknowledge the participants and investigators of the FinnGen study. We thank Pietro Della Briotta Parolo for calculating polygenic risk scores in FinnGen.

Patients and control subjects in FinnGen provided informed consent for biobank research, based on the Finnish Biobank Act. Alternatively, separate research cohorts, collected prior the Finnish Biobank Act came into effect (in September 2013) and start of FinnGen (August 2017), were collected based on study-specific consents and later transferred to the Finnish biobanks after approval by Fimea (Finnish Medicines Agency), the National Supervisory Authority for Welfare and Health. Recruitment protocols followed the biobank protocols approved by Fimea. The Coordinating Ethics Committee of the Hospital District of Helsinki and Uusimaa (HUS) statement number for the FinnGen study is Nr HUS/990/2017.

The FinnGen study is approved by Finnish Institute for Health and Welfare (permit numbers: THL/2031/6.02.00/2017, THL/1101/5.05.00/2017, THL/341/6.02.00/2018, THL/2222/6.02.00/2018, THL/283/6.02.00/2019, THL/1721/5.05.00/2019 and THL/1524/5.05.00/2020), Digital and population data service agency (permit numbers: VRK43431/2017-3, VRK/6909/2018-3, VRK/4415/2019-3), the Social Insurance Institution (permit numbers: KELA 58/522/2017, KELA 131/522/2018, KELA 70/522/2019, KELA 98/522/2019, KELA 134/522/2019, KELA 138/522/2019, KELA 2/522/2020, KELA 16/522/2020), Findata permit numbers THL/2364/14.02/2020, THL/4055/14.06.00/2020,,THL/3433/14.06.00/2020, THL/4432/14.06/2020, THL/5189/14.06/2020, THL/5894/14.06.00/2020, THL/6619/14.06.00/2020, THL/209/14.06.00/2021, THL/688/14.06.00/2021, THL/1284/14.06.00/2021, THL/1965/14.06.00/2021, THL/5546/14.02.00/2020, THL/2658/14.06.00/2021, THL/4235/14.06.00/2021 and Statistics Finland (permit numbers: TK-53-1041-17 and TK/143/07.03.00/2020 (earlier TK-53-90-20) TK/1735/07.03.00/2021). The Biobank Access Decisions for FinnGen samples and data utilized in FinnGen Data Freeze 8 include: THL Biobank BB2017_55, BB2017_111, BB2018_19, BB_2018_34, BB_2018_67, BB2018_71, BB2019_7, BB2019_8, BB2019_26, BB2020_1, Finnish Red Cross Blood Service Biobank 7.12.2017, Helsinki Biobank HUS/359/2017, Auria Biobank AB17-5154 and amendment #1 (August 17 2020), AB20-5926 and amendment #1 (April 23 2020), Biobank Borealis of Northern Finland_2017_1013, Biobank of Eastern Finland 1186/2018 and amendment 22 § /2020, Finnish Clinical Biobank Tampere MH0004 and amendments (21.02.2020 & 06.10.2020), Central Finland Biobank 1-2017, and Terveystalo Biobank STB 2018001. All Finnish Biobanks are members of BBMRI.fi infrastructure (www.bbmri.fi). Finnish Biobank Cooperative –FINBB (https://finbb.fi/) is the coordinator of BBMRI-ERIC operations in Finland.

## Estonian biobank

We want to acknowledge the participants and investigators of the Estonian biobank (EstBB). The EstBB is a population-based biobank managed by the Institute of Genomics at the University of Tartu. It currently contains genotype data and health information for more than 200,000 participants, representing almost 20% of Estonia’s adult population. All participants have provided broad written consent that covers the provision of samples for future research use along with the acquisition of electronic health records from national registries and databases. The activities of the EstBB are regulated by the Human Genes Research Act, which was adopted in 2000 specifically for the operations of the EstBB. Individual level data analysis in the EstBB was carried out under ethical approval 1.1-12/624 from the Estonian Committee on Bioethics and Human Research (Estonian Ministry of Social Affairs), using data according to release application 6-7/GI/11577 from the Estonian Biobank. Data analysis was carried out in part in the High-Performance Computing Center of University of Tartu.

## Bio*Me* biobank

We want to acknowledge the participants and investigators of the Bio*Me* cohort. Founded in September 2007, Bio*Me* is a biobank that links genetic and EMR data for more than 50,000 individuals from diverse ancestral and cultural backgrounds recruited primarily in ambulatory care settings in the Mount Sinai Health System (MSHS) in New York City. The current study was approved by the Icahn School of Medicine at Mount Sinai Institutional Review Board (IRB; approval STUDY-19-00951). All study participants provided written informed consent.

## Author contributions

H.O.H. conceptualized the study, curated and analyzed FinnGen data and wrote the paper. F.D.P. and J.I.D.O. performed replication analyses in the Estonian biobank and Bio*Me* biobank, respectively. J.W. performed analyses with FinnGen data. K.L., R.M. R.K., A.P. provided resources. R.K. provided and oversaw clinical interpretation of the data and results. M.J.D. supervised analytical aspects of the study. All authors read and approved the manuscript.

## Data availability

Individual level data in this study are not publicly available due to legal and privacy limitations, but they can be accessed through individual participating biobanks. The FinnGen data may be accessed through Finnish Biobanks’ FinBB portal (www.finbb.fi). Researchers interested in Estonian Biobank can request access at https://www.geenivaramu.ee/en/access-biobank. For access to data from BioMe biobank, please read here (https://icahn.mssm.edu/research/ipm/programs/biome-biobank). For questions, please reach out to.

## Methods

### Data and definition of epilepsy cases and controls

Here, we define epilepsy case and control status from detailed longitudinal EHR of the FinnGen project ^26^ using data freeze R8 as a main cohort (n=356,036) and Estonian biobank ^27^ as an additional validation cohort (n=210,382). We use phenotype data derived from official state registries. These include 7203 individuals with epilepsy ICD codes, 1,136,442 ASM purchases and 11,306 ASM reimbursements of ATC codes N03A*. We list an overview of case definitions and numbers in Supplementary Table 1. 95.8% of individuals with ≥ 2 generalized seizure ICD codes and 95.6% of individuals with ≥ 2 focal seizure ICD codes purchased ≥ 2 ASMs, while only 14.2% of individuals without epilepsy diagnoses purchased ≥ 2 ASMs (see Supplementary Figure 1). This cross-validates our EHR data.

Reimbursement rights for epilepsy are derived from the Social Insurance Institution of Finland (KELA), Finland’s national authority. All persons with newly diagnosed epilepsy are eligible for ASM reimbursement, which is also routinely applied for, necessitating a detailed statement by a neurologist and investigations at a specialist clinic. The statement is checked and approved by specialist physicians at the reimbursement institution KELA before the right is granted. Epilepsy diagnoses in Finland are made according to national guidelines, which are updated according to ILAE epilepsy definitions.

We thus chose following criteria to define ***GGE***:

– at least two ICD codes of G40.3 or corresponding ICD9 codes (Supplementary Table 2) and at least two purchases of ASMs (as defined by N03* ATC codes).

We chose following criteria to define ***NAFE***:

– at least two ICD codes of G40.0, G40.1, G40.2 or corresponding ICD9 diagnoses (Supplementary Table 2) and at least two purchases of ASMs.

– excluded possible structural aetiology of focal seizures such as stroke, brain tumour, CNS infection and CNS injury (for ICD codes see Supplementary Table 2). Here, we only excluded individuals if they had their first seizure event within one year after the brain-related potential epileptogenic event.

For 606 individuals with both focal and generalized epilepsy codes we applied following additional criteria for a GGE diagnosis:

– more generalized than focal epilepsy codes AND

– most frequent ICD code is a generalized epilepsy code AND

– no reimbursement category of focal epilepsy

We used the same criteria vice versa to define NAFE among individuals with focal and generalized epilepsy codes.

We defined ***idiopathic generalized epilepsy*** (IGE) according to ILAE ^29,44^ by at least two ICD codes of 40.33 (Childhood Absence Epilepsy), 40.34 (Generalized Tonic–Clonic Seizures Alone, here using the ICD Code of the formerly known term Generalized Tonic–Clonic Seizures on Awakening) 40.35 (Juvenile Absence Epilepsy), 40.36 (Juvenile Myoclonic Epilepsy). See Supplementary Figure 2 for age at first diagnosis.

We used individuals without epilepsy related diagnoses as controls. We excluded individuals who purchased ASMs from the control group.

For the analysis of GGE incidence following an unspecified seizure event, we used the same diagnosis of GGE as described above. We defined an unspecified seizure event with an ICD code of R56.8/7803A (‘unspecified convulsions’). From the group with a single unspecified seizure event we excluded individuals

– with any other epilepsy-related diagnoses (G40/G41 ICD codes) AND

– who purchased or reimbursed ASMs within two years before up to 10 years after event AND

– who were at any time diagnosed with alcohol-related ICD codes OR who had multiple unspecified seizure events (to exclude potential alcohol withdrawal seizures).

When individuals had 2 seizure diagnoses on the same day we counted them as one seizure event as they most likely represent two labels of the same event. When the 2 seizure diagnoses had discordant ICD labels we labeled them according to the most specific ICD code. (As an example, individuals with diagnoses of unspecified seizure and generalized epilepsy on initial presentation would be classified as diagnosed with ‘generalized epilepsy’ on initial presentation.)

We defined epilepsy cases similarly in the validation cohort Estonian biobank, with the only exception that instead of using the reimbursement data to differentiate between NAFE and GGE in individuals who had both focal and generalized epilepsy codes, we used prescription data. Specifically, we excluded individuals as GGE cases if they had any ASM prescriptions that listed NAFE as a reason for the prescription and vice versa. We performed the unspecified seizure event analysis only in FinnGen where we had a sufficient sample size.

### Calculation of polygenic risk scores

We calculated epilepsy PRS with the method PRS-CS ^45^. Here, we used the ILAE GWAS 2018 ^12^ summary statistics as discovery data, i.e. to determine which genetic variants increase or decrease epilepsy risk. We constructed separate focal PRS (PRS_NAFE_) and generalized epilepsy PRS (PRS_GGE_). The cohorts in our study did not overlap with the ILAE GWAS cohort. We applied the PRS-CS-auto algorithm to infer posterior effect sizes for the variants for PRS calculation. PRS-CS-auto learns the model’s global scaling parameter ϕ from the data. We used data from the 1000 Genomes ^46^ as a reference panel for linkage disequilibrium. We then weighted and summed >835k genetic variants that confer either risk for or protection from epilepsy with individually small effects into a single epilepsy PRS per individual using the PLINK –-score command ^47^. The PRS-CS pipeline in FinnGen is described in more details at https://github.com/FINNGEN/CS-PRS-pipeline.

In FinnGen and EstBB, we restricted our analysis to individuals with European ancestry, while we additionally included African and American continental ancestry groups in the Bio*Me* cohort (Supplementary Note). We inferred population labels based on principle component analysis of the genotype data as described previously ^26,27^.

### Statistical analyses

We used the R programming language for all statistical analyses. In all analyses including PRSs namely logistic regression, survival analyses and concordance index calculations we included following covariates: the first 10 principal components of genetic markers (10 PCs) as a proxy for population substructure and ancestry, genotyping batch (only in the FinnGen cohort), sex, birth year, age at last follow up. For analyses that included only individuals with seizures, we included age at first epilepsy diagnosis as covariate instead of age at last follow-up. We tested PRS_GGE_ or PRS_NAFE_ as indicated in the results of the manuscript. We did not exclude individuals that were related as we found in extensive sensitivity analyses that this did not influence PRS effect on disease ^48^.

In the PheWas, we defined independent diseases when for any disease category not more than 40% of affected individuals are listed in any other disease category.

We performed survival analyses using the Cox Proportional-Hazards model (Cox-PH) ^30^. Follow-up started at birth and ended at the age of first epilepsy diagnosis (for individuals with epilepsy), age at last record available in the EHR or death, depending on what happened first. We also performed survival analyses in individuals with an unspecified seizure. Here, follow-up started at the age of the unspecified seizure and ended at the age of first epilepsy diagnosis, age at last record available in the EHR, death or after 10 years, depending on what happened first. We tested for sex differences by including an interaction term of PRS x sex in the Cox-PH model. We used the first 10 PCs, genotyping batch, sex, birth year and age at last follow up as covariates in all survival analyses. We excluded sex in sex-specific survival analyses. We included age at first unspecified seizure in survival analyses of individuals with an unspecified seizure.

