## Supplementary Information for "Polygenic risk scores as a marker for epilepsy risk across lifetime and after unspecified seizure events"

### Supplementary Figures

**Supplementary Figure 1. Antiseizure medication (ASM) purchases.** The three barplots show ASM purchases of individuals with at least two epilepsy diagnosis codes of GGE or NAFE, respectively, and without epilepsy diagnosis codes (‘control’). Counts are grouped for individuals who made 0 (blue), 1 (red) and ≥2 (yellow) ASM purchases, respectively.

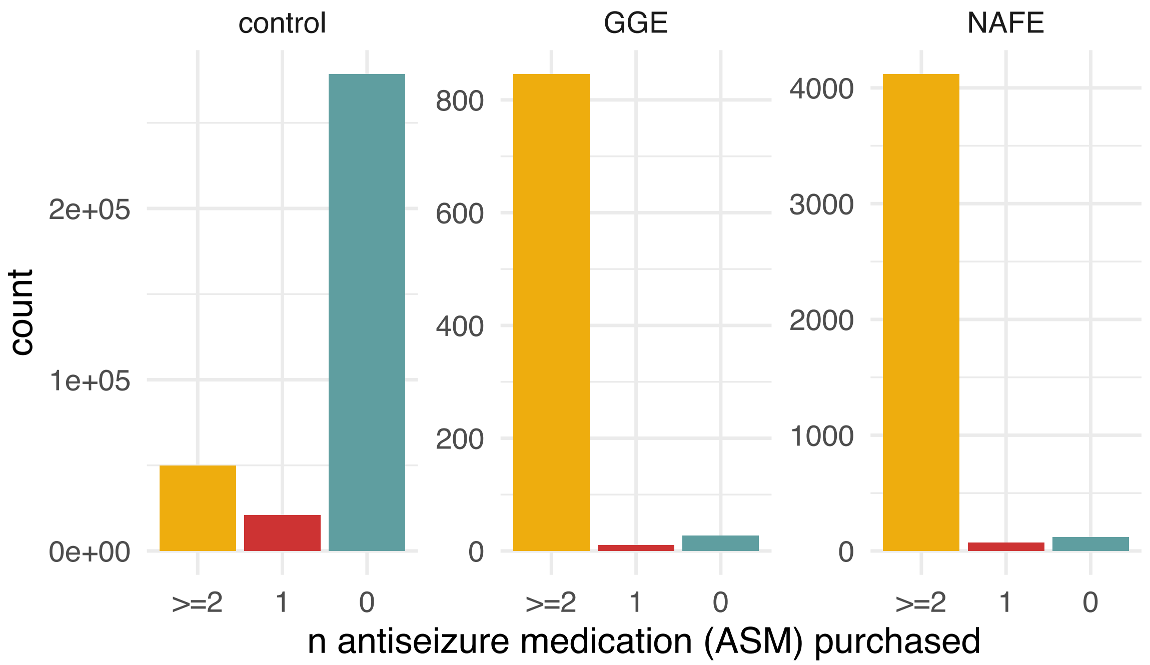

**Supplementary Figure 2. Age at IGE diagnosis.** Age at first diagnosis of individuals with at least 2 diagnoses of idiopathic generalized epilepsy diagnoses (=IGE) [1]. The four panels show age at diagnosis of individuals with most frequent ICD codes of Childhood Absence Epilepsy (G40.33, n=19), Generalized Tonic–Clonic Seizures on Awakening (*now:* Generalized Tonic–Clonic Seizures Alone) (G40.34, n=8), Juvenile Absence Epilepsy (G40.35, n=22) Juvenile Myoclonic Epilepsy (G40.36, n=82).

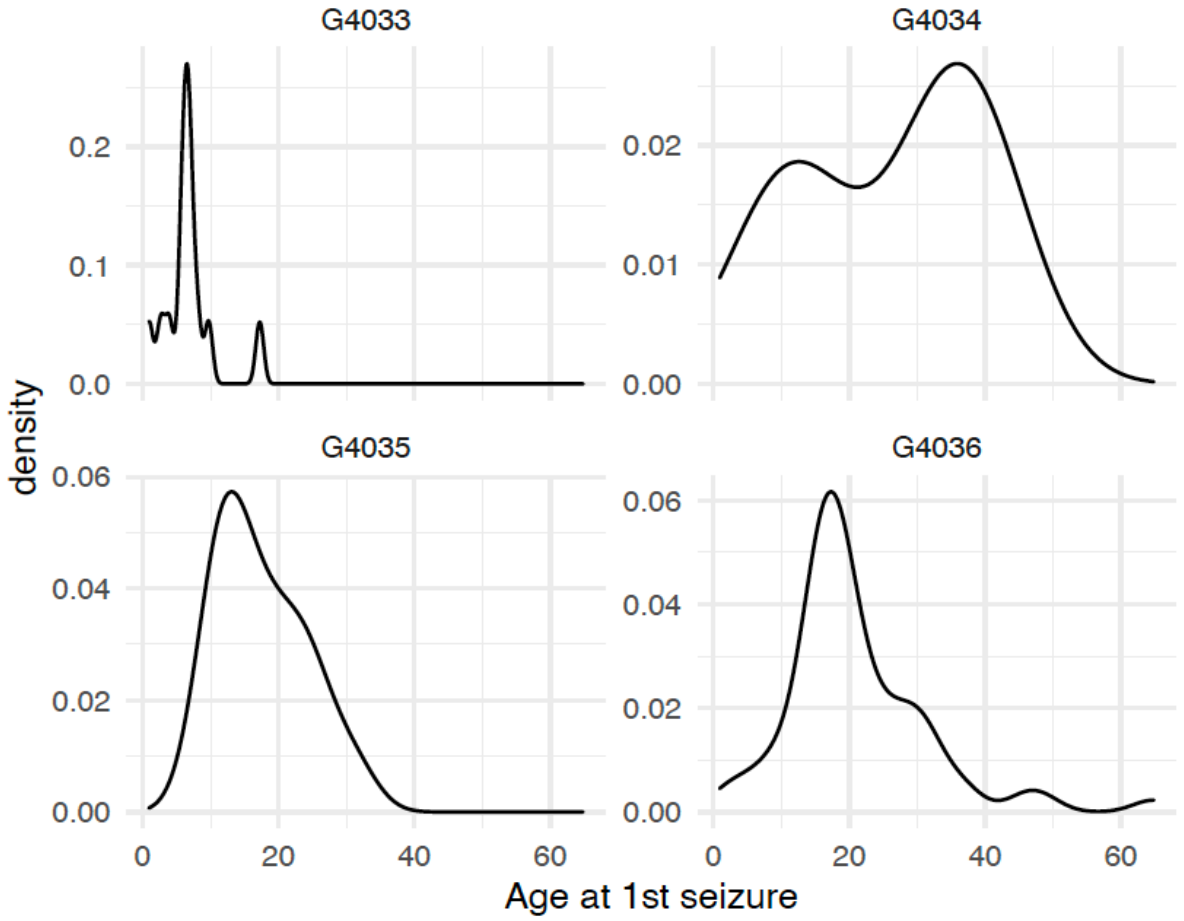

**Supplementary Figure 4**. **Epilepsy PRS as a marker for epilepsy risk across lifetime in 164,621 individuals from the Estonian biobank.** In panel A, we investigate the effect of $PRS_{GGE}$ on GGE across lifetime; in panel B, we investigate the effect of $PRS_{NAFE}$ on NAFE across lifetime. In each panel, on the left are density curves that display how samples are partitioned into six bins of PRS standard deviations. Survival curves in the middle give the cumulative epilepsy incidence (y-axis) across time (x-axis [years]) stratified for epilepsy PRS bins. The rightmost figures show epilepsy risk of each epilepsy PRS bin compared with the rest of the cohort (forest plots). Here, the point estimates represent hazard ratios, error bars show the 95%-confidence intervals.

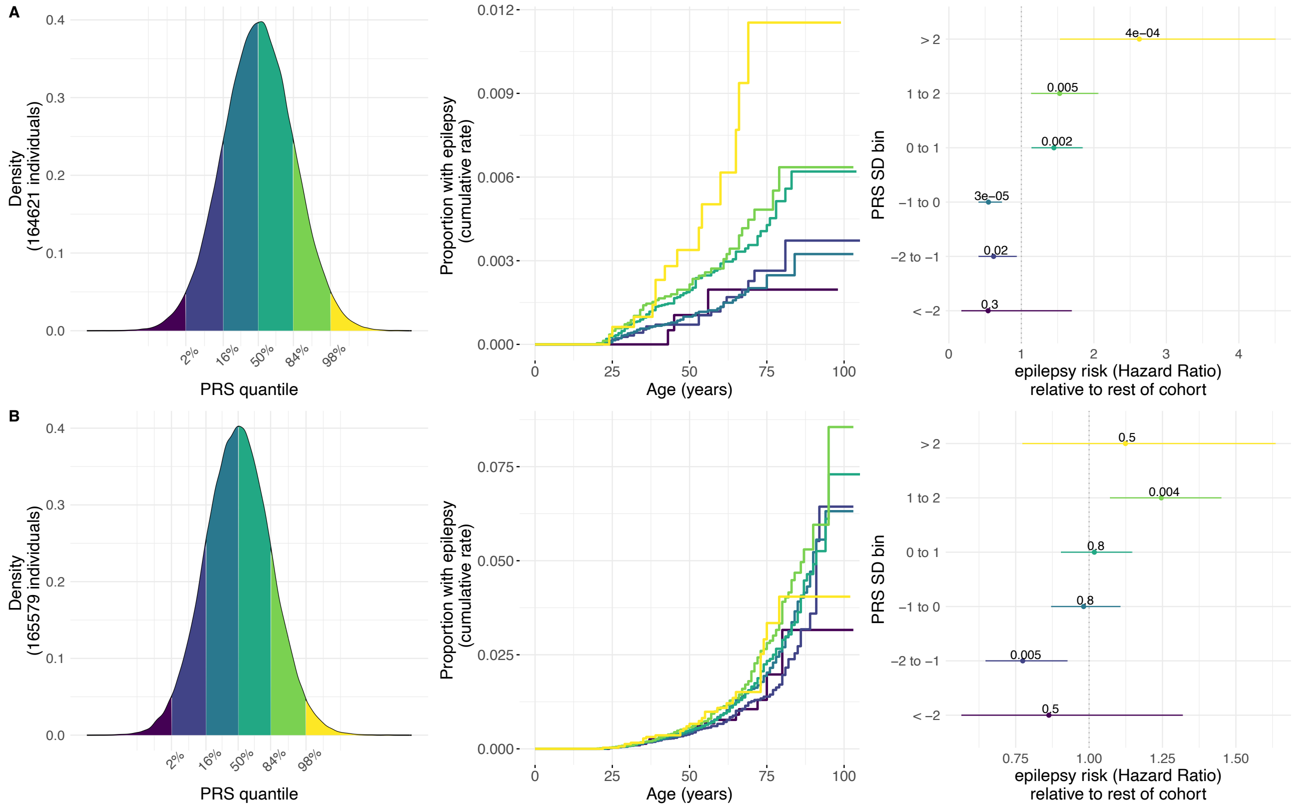

**Supplementary Figure 5**. **Effect of epilepsy PRS$_{GGE}$ on lifetime GGE incidence in 18,152 individuals with European (EUR), African (AFR) and American (AMR) ancestries from the Bio*Me* cohort (Mount Sinai Hospital, New York).** In each panel, survival curves show the cumulative lifetime epilepsy incidence (y-axis) across age (x-axis [years]) stratified for bins of top (>75$^{th}$ percentile, blue), joined two middle (25$^{th}$-75$^{th}$ percentile, green) and bottom quartiles (<25$^{th}$ percentile, red) of epilepsy PRS$_{GGE}$. Different panels show different genetic ancestry groups. A) EUR ancestry, B) AMR ancestry, C) AFR ancestry.

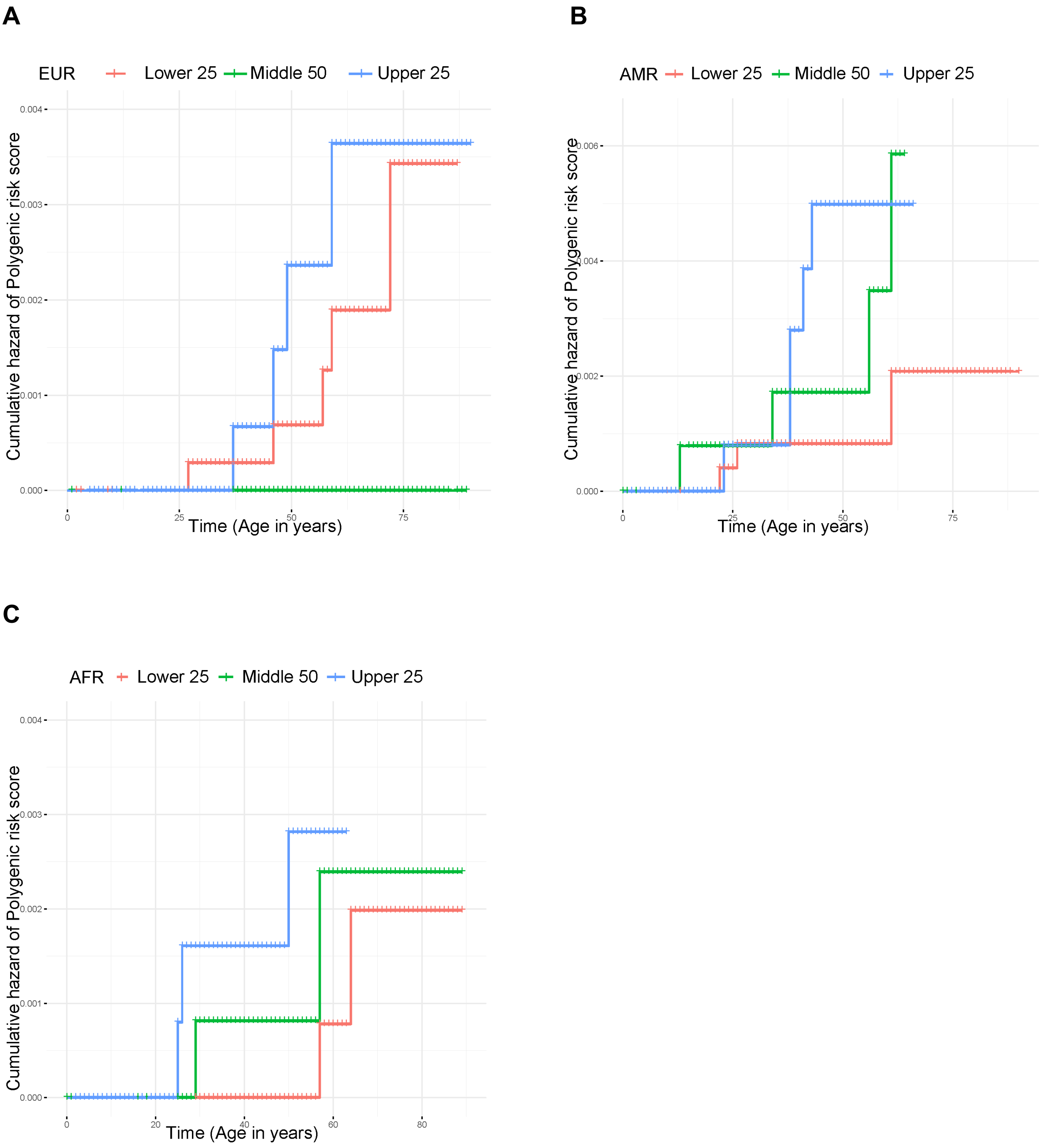

**Supplementary Figure 6**. **Sex-specific differences on the effect of $PRS_{GGE}$ on epilepsy prevalence across lifetime.** Panels *Female* and *Male* indicate that the analysis was performed in only-female (n=144,630) and only-male (n=119,054) individuals, respectively. In both panels, on the left are density curves that display how samples are partitioned into six bins of PRS standard deviations. Survival curves in the middle give the cumulative epilepsy incidence (y-axis) across time (x-axis [years]) stratified for epilepsy PRS bins. The rightmost figures show epilepsy risk of each epilepsy PRS bin compared with the rest of the cohort (forest plots). Here, the point estimates represent hazard ratios, error bars show the 95%-confidence intervals.

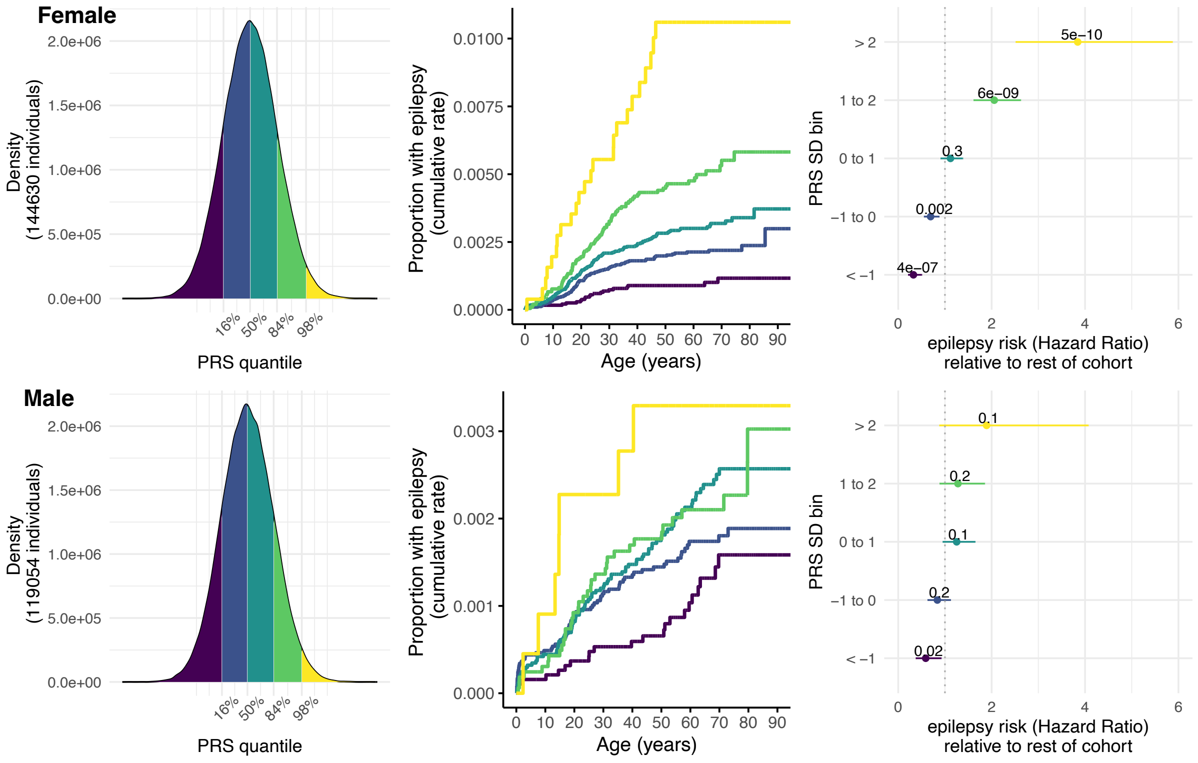

**Supplementary Figure 7. Genetic correlations between NAFE and the 18 disease phenotypes that were significantly associated with $PRS_{NAFE}$.** Disease phenotypes, sorted by clinical field, are shown on the x-axis. The genetic correlation coefficient ($r_g$) is visualized with a color scale ranging from -1 (blue) to 1 (red). Corresponding p-values of the correlations are shown inside the boxes. Genetic correlations were calculated with LD score regression [6]. The GWAS summary statistics were taken from FinnGen, release 10 (n = 430,897).

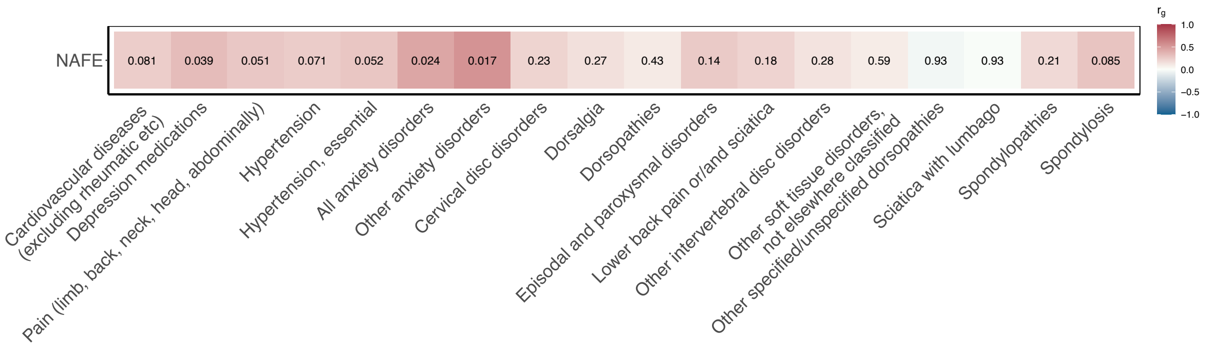

### Supplementary Tables

#### Supplementary Table 1. Definitions of epilepsy cases and controls

| Category | n R8 | Definition |
|---|---|---|
| Control | 273,974 | exclude ASM purchases/reimbursement |
| Focal | 3,127 | ≥ 2x G40.0-G40.2; ≥2 ASM purchases, exclude head injury, infection, tumor or stroke < 1 year before epilepsy (see Supplementary Table 2) |
| Generalized | 622 | ≥ 2x G40.3, ≥2 ASM purchases |
| Other epilepsy diagnoses | 3,242 | ≥ 1 x G40 or G41 |
| Single unspecified seizure | 1,806 | R56.8 or 7803A; exclude individuals with ASM purchase/ reimb. within 10 years after and 2 years before seizure event |
| Possible alcohol withdrawal seizures | 3,655 | Multiple unspecified seizures/epilepsy OR<br>alcohol-related seizures OR<br>unspecified seizures/epilepsy & alcohol related diagnoses |

**Supplementary Table 2. Definitions of epilepsy ICD codes**

| Code | Category | ICD | Comment |
|---|---|---|---|
| **Unspecified seizure diagnosis codes** | | | |
| R56.8 | unspecified | 10 | |
| 7803A | unspecified | 9 | |
| **Focal and general epilepsy diagnosis codes** | | | |
| G40.3 | generalized | 10 | |
| G40.30 | generalized | 10 | |
| G40.31 | generalized | 10 | |
| G40.33 | generalized | 10 | IGE (Childhood Absence Epilepsy) |
| G40.34 | generalized | 10 | IGE (Generalized Tonic–Clonic Seizures on Awakening) |
| G40.35 | generalized | 10 | IGE (Juvenile absence epilepsy) |
| G40.36 | generalized | 10 | IGE (Juvenile myoclonic epilepsy) |
| G40.39 | generalized | 10 | |
| 3450A | generalized | 9 | |
| 3451A | generalized | 9 | |
| 3451 | generalized | 9 | |
| 3452A | generalized | 9 | |
| 3453A | generalized | 9 | |
| G40.0 | focal | 10 | |
| G40.00 | focal | 10 | |
| G40.01 | focal | 10 | |
| G40.09 | focal | 10 | |
| G40.1 | focal | 10 | |
| G40.10 | focal | 10 | |
| G40.11 | focal | 10 | |
| G40.12 | focal | 10 | |
| G40.19 | focal | 10 | |
| G40.2 | focal | 10 | |
| G40.20 | focal | 10 | |
| G40.21 | focal | 10 | |
| G40.22 | focal | 10 | |
| G40.29 | focal | 10 | |
| 3454 | focal | 9 | |
| 3454A | focal | 9 | |
| 3454B | focal | 9 | |
| 3454X | focal | 9 | |
| 3455A | focal | 9 | |

| | | | |
|---|---|---|---|
| **Epilepsy syndromes, status epilepticus and other epilepsy diagnosis codes** | | | |
| G41 | unclassified epilepsy | 10 | Status epilepticus |
| G41.0 | unclassified epilepsy | 10 | Status epilepticus |
| G41.1 | unclassified epilepsy | 10 | Status epilepticus |
| G41.2 | unclassified epilepsy | 10 | Status epilepticus |
| G41.8 | unclassified epilepsy | 10 | Status epilepticus |
| G41.9 | unclassified epilepsy | 10 | Status epilepticus |
| G40.50 | unclassified epilepsy | 10 | Status epilepticus |
| 3457A | unclassified epilepsy | 9 | Status epilepticus |
| 3457B | unclassified epilepsy | 9 | Status epilepticus |
| 3457X | unclassified epilepsy | 9 | Status epilepticus |
| G40 | unclassified epilepsy | 10 | |
| G40.6 | unclassified epilepsy | 10 | |
| G40.7 | unclassified epilepsy | 10 | |
| G40.8 | unclassified epilepsy | 10 | |
| G40.4 | unclassified epilepsy | 10 | |
| G40.5 | unclassified epilepsy | 10 | DEE |
| G40.59 | unclassified epilepsy | 10 | DEE |
| G40.80 | unclassified epilepsy | 10 | DEE |
| G40.89 | unclassified epilepsy | 10 | DEE |
| G40.37 | unclassified epilepsy | 10 | PME |
| 345 | unclassified | 9 | |
| 3456A | unclassified epilepsy | 9 | DEE |
| 3456B | unclassified epilepsy | 9 | GEFSplus |
| 3458X | unclassified epilepsy | 9 | |
| G40.9 | unclassified epilepsy | 10 | |
| 3459X | unclassified epilepsy | 9 | |
| **Exclusion ICD codes** | | | |
| G40.51 | alcohol related seizure | 10 | |
| C71 | symptomatic focal epilepsy | 10 | Malignant neoplasm of brain |
| 191 | symptomatic focal epilepsy | 9 | Malignant neoplasm of brain |
| 191 | symptomatic focal epilepsy | 8 | Malignant neoplasm of brain |
| C70 | symptomatic focal epilepsy | 10 | Malignant neoplasm of meninges |
| 1921\|1923 | symptomatic focal epilepsy | 9 | Malignant neoplasm of meninges |
| 192[1-2] | symptomatic focal epilepsy | 8 | Malignant neoplasm of meninges |
| C72 | symptomatic focal epilepsy | 10 | Malignant neoplasm of spinal cord, cranial nerves and other parts of central nervous system |
| 192 | symptomatic focal epilepsy | 9 | Malignant neoplasm of spinal cord, cranial nerves and other parts of central nervous system |
| 192 | symptomatic focal epilepsy | 8 | Malignant neoplasm of spinal cord, cranial nerves and other parts of central nervous system |

| | | | |
|---|---|---|---|
| I6[0-4]\|G45 | symptomatic focal epilepsy | 10 | stroke |
| 430\|4330A\|4331A\|4339A\|4340A\|4341A\|4349A\|436\|435 | symptomatic focal epilepsy | 9 | stroke |
| 430\|431\|433\|434\|435\|436 | symptomatic focal epilepsy | 8 | stroke |
| S00-S09 | symptomatic focal epilepsy | 10 | head injury |
| G00-G09 | symptomatic focal epilepsy | 10 | Inflammatory diseases of the central nervous system |

**Supplementary Table 3. Enrichment of epilepsy cases in individuals with high epilepsy PRS.**

Odds ratios for epilepsy case status were calculated comparing individuals in bins of the top 0.5%, 5% and 20% of epilepsy PRS with the remainder of the cohort as done in [2]. As our GGE cohort size was more modest an additional 2.5% bin was added for GGE. PRS was matched to epilepsy type, i.e. the effect of $PRS_{NAFE}$ on NAFE case status, and the effect of $PRS_{GGE}$ on GGE case status was estimated. The results were very similar compared to clinically defined epilepsy cases previously described in [2] (Table 2) validating our epilepsy phenotyping approach. Method: logistic regression. Covariates: sex, birth year, age at last follow up, first 10 PCs of ancestry, genotyping batch. OR; Odds ratio, CI; confidence interval. Dataset: FinnGen.

| PRS/ phenotype | PRSbin | OR | 5%-CI | 95%-CI | P-value | n cases in bin | n cases total | n controls in bin | n controls total |
|---|---|---|---|---|---|---|---|---|---|
| GGE | 20% | 1.98 | 1.65 | 2.37 | 1.2E-13 | 197 | 622 | 54733 | 273974 |
| GGE | 5% | 2.39 | 1.79 | 3.12 | 6.5E-10 | 69 | 622 | 13772 | 273974 |
| GGE | 2.5% | 3.13 | 2.19 | 4.33 | 4.6E-11 | 44 | 622 | 6905 | 273974 |
| GGE | 0.5% | 2.55 | 1.00 | 5.28 | 2.5E-02 | 8 | 622 | 1365 | 273974 |
| NAFE | 20% | 1.23 | 1.13 | 1.34 | 2.5E-06 | 755 | 3127 | 54436 | 273974 |
| NAFE | 5% | 1.39 | 1.20 | 1.61 | 1.3E-05 | 212 | 3127 | 13672 | 273974 |
| NAFE | 0.5% | 2.07 | 1.18 | 3.34 | 5.8E-03 | 21 | 3127 | 1449 | 273974 |

**Supplementary Table 4. Effect of epilepsy PRS on epilepsy at different ages of epilepsy onset.**

Odds ratios for epilepsy case status were calculated comparing individuals without epilepsy to individuals with epilepsy age at seizure onset in respective age bins of 20 years.
Epilepsy PRS was z-transformed, thus odds ratios are given per SD increase of epilepsy PRS.
Method: logistic regression. Covariates: sex, birth year, age at last follow up, first 10 PCs of ancestry, genotyping batch. OR; Odds ratio, CI; confidence interval. Dataset: FinnGen.

| PRS/ phenotype | age bin (years) | OR | 5%-CI | 95%-CI | P-value | n cases in bin | n controls in bin |
|---|---|---|---|---|---|---|---|
| GGE | 0-20 | 1.61 | 1.42 | 1.82 | 2.05E-13 | 295 | 273974 |
| GGE | 20-40 | 1.59 | 1.37 | 1.85 | 1.10E-09 | 200 | 273974 |
| GGE | 40-60 | 1.49 | 1.19 | 1.87 | 0.00056 | 88 | 273974 |
| GGE | 60-80 | 1.18 | 0.82 | 1.69 | 0.37 | 36 | 273974 |
| NAFE | 0-20 | 1.12 | 1.02 | 1.22 | 0.02 | 589 | 273974 |
| NAFE | 20-40 | 1.15 | 1.06 | 1.24 | 0.00066 | 731 | 273974 |
| NAFE | 40-60 | 1.13 | 1.06 | 1.22 | 0.00039 | 937 | 273974 |
| NAFE | 60-80 | 1.12 | 1.04 | 1.21 | 0.0036 | 774 | 273974 |
| NAFE | 80-100 | 1.12 | 0.90 | 1.40 | 0.31 | 94 | 273974 |

### Supplementary Notes

#### Replication in Bio*Me* cohort

##### *Summary*

We sought to replicate our analysis in other ancestry groups. We thus tested association of $PRS_{GGE}$ with GGE in individuals with more diverse ancestries from the Bio*Me* cohort. Due to power, we restricted our analysis to populations of European (n= 7329), African (n= 5587) and American (n= 5236) genetic ancestry, respectively. In Supplementary Figure 5 we show survival curves stratifying the cumulative incidence of GGE according to polygenic risk quartiles across the respective genetic ancestries.

##### *Methods*

We replicated the analysis in the Bio*Me* cohort. Same as in the other two cohorts, $PRS_{GGE}$ was calculated using summary statistics of the International League Against Epilepsy Consortium on Complex Epilepsies [3]. Epilepsy cases were derived from participants' EHR, using ICD-9 and -10 codes as described in Supplementary Table 2. Genetic ancestries of Bio*Me* participants were inferred using Principal Component Analysis (PCA) of genotypes using a random forest model. The model was trained using population labels from reference data of the 1000 genomes project, namely European, Admixed American, East Asian, South Asian, and African continental populations. Weight files for each continental ancestry group in the BioMe cohort were computed using the tool PRS-CS [4] and reference data of the 1000 genomes project, and PRSs of individuals calculated using PLINK [5]. The $PRS_{GGE}$ was z-transformed to calculate HR per SD of $PRS_{GGE}$ across lifetime with a Cox-proportional Hazards model.

##### *Results*

| Epilepsy/PRS | N epilepsy | N total | Ancestry | HR per SD $PRS_{GGE}$ (95%- CI) | p-value |
|---|---|---|---|---|---|
| GGE | 10 | 7329 | European | 1.5 (0.8 - 2.8) | 0.16 |
| GGE | 8 | 5587 | American | 1.0 (0.6 - 1.7) | 0.95 |
| GGE | 14 | 5236 | African | 1.5 (0.7 - 3.1) | 0.26 |
| GGE | 37 | 19255 | European, African, American, South Asian, East Asian, | 1.3 (0.9 - 1.7) | 0.16 |

**Effects of PRS on epilepsy risk in different cohorts.** Epilepsy risks are given as hazard ratios (HR) for epilepsy per SD increase of epilepsy PRS. Method: Cox proportional hazards model. CI; confidence interval. Results are shown for ancestry groups with ≥ 3 epilepsy cases as well as a joint analysis of all individuals.
