## Supplementary material for "Polygenic risk scores as a marker for epilepsy risk across lifetime and after unspecified seizure events": Estonian biobank banner

### Estonian Biobank research team

| Full Name | Affiliation |
|---|---|
| | Institute of Genomics, University of Tartu, Estonia |
| Andres Metspalu | The Institute of Molecular and Cell Biology, University of Tartu, Estonia |
| Lili Milani | Institute of Genomics, University of Tartu, Estonia |
| Tõnu Esko | Institute of Genomics, University of Tartu, Estonia |
| Reedik Mägi | Institute of Genomics, University of Tartu, Estonia |
| Mari Nelis | Core Facility of Genomics, Institute of Genomics, University of Tartu, Estonia |
| Georgi Hudjashov | Institute of Genomics, University of Tartu, Estonia |

| E-mail | Role 1 | Role 2 |
|---|---|---|
| | sample collection | Head of the EstBB cohort |
| | | |
| | | |
| | QC of the data | |
| | Sample genotyping | |
| | | |
